# High-Resolution Variant Profiling of CAH-Associated Genes Using a Long-Read Sequencing Assay

**DOI:** 10.1101/2025.10.24.25338696

**Authors:** Shanshan Gu, Guoming Chu, Ruining Cai, Xiangzhong Sun, Qinglong Lu, Wanqing Han, Senwen Deng, Xiaohua Wang, Jiale Xiang, Rong He

## Abstract

**Background:** Congenital adrenal hyperplasia (CAH) is a group of autosomal recessive disorders primarily caused by mutations in *CYP21A2* and *CYP11B1*. However, accurate genetic diagnosis remains challenging due to the high sequence homology between these genes and their corresponding homologs (*CYP21A1P, CYP11B2*).

**Methods:** We developed NanoCAH (Nanopore-based Comprehensive Analysis of CAH), a targeted long-read sequencing assay based on the Cyclone nanopore platform, CycloneSEQ G100-ER. This approach uses gene-specific long-range PCR to amplify *CYP21A2* and *CYP11B1*, enabling detection of all classes of variants including single nucleotide variants (SNVs), copy number variants (CNVs), and gene conversions.

**Results:** A total of 59 samples were analyzed by NanoCAH, including 26 singletons and 11 trios. Among these samples, NanoCAH successfully detected all 85 variants previously identified by conventional MLPA and Sanger sequencing (100%, 85/85), including SNVs/Indels, deletions and duplications. Importantly, seven Exon 8 deletions and one Exon 8 duplication undetectable by MLPA were identified by NanoCAH assay. In addition, NanoCAH resolved a novel heterozygous 111-bp in-exon tandem duplication in *CYP21A2* (c.65_175dup). NanoCAH further accurately resolved haplotypes in twenty cases without parental data, identifying 18 variants in trans and 2 in cis. These results demonstrate NanoCAH’s robust capacity to detect complex variant types and provide precise haplotype resolution in a single assay.

**Conclusions:** NanoCAH provides an accurate, cost-effective, and scalable solution for comprehensive CAH genotyping. Its ability to detect complex variant types and resolve haplotypes in a single assay highlights its potential as a first-line diagnostic tool in clinical genetics.

## INTRODUCTION

Congenital adrenal hyperplasia (CAH) encompasses a group of inherited genetic disorders that disrupt cortisol biosynthesis, primarily due to reduced enzyme activity (1). The most common form of CAH is caused by deficiencies in steroid 21-hydroxylase (21OH), resulting from mutations in the *CYP21A2* gene. 21OH deficiency CAH accounts for 90-95% of CAH. The second most common form of CAH after *CYP21A2* is 11-beta-hydroxylase (11β-OH) deficiency CAH (5-8% of all CAH cases), caused by variants from *CYP11B1* (2). In addition to these, rare genes include *CYP17A1, HSD3B2, StAR*, *CYP11A1*, and *POR* can also cause CAH (3). Of them, *CYP21A2* and *CYP11B1* are two genes having highly homologous sequences, posing significant challenges for the molecular detection (4).

*CYP21A2*, encoding 21-hydroxylase, locates on chromosome 6p21.3. The non-functional *CYP21A1P* pseudogene, locating at approximately 30 kb distance, shares approximately 98% nucleotide sequence homology with *CYP21A2*. *CYP21*, together with other genes (serine/threonine kinase (RP), complement (C4), and tenascin (TNX)) form the RCCX module (RP-C4-CYP21-TNX). The high homology between the functional genes (*RP1, CYP21A2*, and *TNXB*) and the corresponding pseudogene (*RP2, CYP21A1P,* and *TNXA*) increases the incidence of gene conversion and the formation of chimeric genes (3). Currently, over 200 pathogenic variants of *CYP21A2* gene have been identified in humans, of which approximately 75% are due to the microconversion of the *CYP21A1P* pseudogene mutation, 20% to 25% are large deletions or chimeric genes, and 1% to 2% are *de novo* mutations.

In contrast to *CYP21A2* and its nonfunctional *CYP21A1P* pseudogene, *CYP11B1* and *CYP11B2* are both active and do not have a pseudogene (2). The two genes are 40-kb apart, each comprising nine exons and mapped to chromosome 8q21-22, which have distinct functions in cortisol and aldosterone synthesis, respectively. *CYP11B2/CYP11B1* shares 95% of sequence similarity within their nine exons and approximately 90% homology in their introns (5).

Due to the highly homologous nature of these sequences, short-read sequencing technologies, which typically generate 150 bp paired-end reads (300 bp in total), have inherent limitations in identifying variants in these regions (6). Traditionally, multiplex ligation-dependent probe amplification (MLPA) and Sanger sequencing have been the primary methods for detecting copy number variants (CNVs) and single nucleotide variants (SNVs) and small insertions and deletions (Indels) in *CYP* genes, respectively (7, 8). More recently, long-read sequencing (LRS) using the PacBio platform has been reported as a reliable approach for detecting *CYP21A2*-related CAH (9, 10). To date, *CYP11B1* has not yet been validated using this method. Additionally, the nanopore sequencing technique, another major LRS approach, has not been demonstrated as effective for CAH detection.

In this study, we developed NanoCAH, a targeted LRS assay on the nanopore sequencing platform that enables comprehensive analysis of variant types, including CNVs, SNVs/Indels, and gene conversions in CAH-related genes (*CYP21A2* and *CYP11B1*). Tested on 59 samples, the assay showed complete agreement with variants identified by MLPA and Sanger sequencing, while additionally detecting variants beyond their coverage. The LRS based assay identified novel deletions/duplications uncovered by MLPA, and accurately resolved compound heterozygous variants through haplotype phasing. Our results highlighted the advantage of LRS method and its clinical applications on the nanopore sequencing platform.

## MATERIALS AND METHODS

### Samples and study design

A total of 59 genomic DNA (gDNA) samples were analyzed. Among them, 33 samples were derived from 11 parent-offspring trios, while the remaining 26 were singleton samples, including both affected individuals and carriers. 19 Samples were identified through MLPA and sanger at the clinical laboratory of Beijing Genomics Institution, China, and other 40 Samples were identified at Shengjing Hospital of China Medical University. All 59 samples had at least one pathogenic variant confirmed by SALSA MLPA Probemix P050-C1 CAH (MRC-Holland) and Sanger sequencing. The study was approved by the institutional review boards of BGI-IRB25011.

### Long-Read PCR and library Construction

The optimized multiplex long-read PCR was conducted in a 25 μL reaction system containing 25ng of gDNA, 1 μmol/L of a mixture of PCR primers, 12.5 μL of 2× KeyPo SE Master Mix (Vazyme Biotech, China), and nuclease-free water added up to 25 μL. The PCR reaction conditions were as follows: initial denaturation at 94°C for 5 minutes, followed by 35 cycles of denaturation at 98°C for 30 seconds, annealing and extension at 68°C for 8 minutes, and a final extension at 68°C for 10 minutes before storing the PCR products at 4°C. The long-range PCR products were purified using 0.5× volume of VAHTS DNA Clean Beads (Vazyme Biotech, China).

Library construction was performed using the CycloneSEQ (Cyclone) kit, according to the kit manual (11). Briefly, 500 ng of purified DNA products were end-repaired using EA Enzyme (CycloneSEQ, China). The end-repaired product was then purified using 0.4× volume of DNA clean beads and eluted with nuclease-free water. Next, the end-repaired and A-tailed product was ligated with barcodes. All ligated products were then mixed, purified using 0.4× volume of DNA clean beads, and eluted with nuclease-free water. Finally, the purified products were ligated to adapters using the Cyclone ligation module, purified with 0.4× volume of DNA clean beads, and eluted with Elution Buffer. The concentration of the final ligation product was measured, and the product was used as the sequencing library.

### CycloneSEQ Long-Read Nanopore Sequencing

Firstly, CycloneSEQ G100-ER sequencer was started, and the chip was replaced using the chip replacement buffer. Chip quality control was then performed to confirm the number of active sequencing nanopores. According to the Cyclone sequencing kit manual, sequencing reagents and anchoring reagents were used to load 400 ng of the sequencing library onto the Cyclone sequencing chip. Sequencing was then initiated, and the basecalling was performed from the raw electrical signal. The entire sequencing run was completed within 48 hours.

### Bioinformatics Analysis

The bioinformatics pipeline comprises two sequential steps. First, we constructed a comprehensive reference genome database for *CYP21A2*, including the identification of differential loci between the functional gene, *CYP21A2*, and its pseudogene, *CYP21A1P.* Second, we implemented an automated pipeline to analyze and annotate the CAH-related genes (*CYP21A2* and *CYP11B1*).

#### 1) Reference database construction

To prepare the database, we first constructed locus-specific reference sequences for both the *CYP21A2* and *CYP21A1P* regions and compiled a list of pseudogene-specific differential sites. Specifically, we extracted the genomic region chr6:32033327-32049480 (hg38) to represent the *CYP21A2-TNXB* locus, and chr6:32000590-32017007 to represent the *CYP21A1P-TNXA* locus. These reference sequences were then used to build a variant comparison database for accurate pseudogene discrimination during downstream analysis. The *CYP21A1P-TNXA* sequence was aligned to the reference *CYP21A2-TNXB* using *Minimap2* (version 2.17-r941) (12) to generate Bam files, which were sorted and indexed using *SAMtools* (version 1.10) (13). Subsequently, SNV calling was performed using *Freebayes* (version 1.3.6) (14), and the results were normalized using *Bcftools* (version 1.10.2) (13) to generate the database of differential sites for pseudogenes. The database includes the positions of each site as well as the corresponding bases of the functional genes and pseudogenes.

#### 2) Automated sample processing

Raw FASTQ data generated by the CycloneSEQ G100-ER sequencer were processed using *Fastp* (v0.23.4) (15). Reads were retained if they were longer than 1400 bp and had a Phred quality score ≥ 7. Filtered reads were classified by primer pairs using *Jasper*, a custom-developed software for primer pair splitting. Reads associated with *CYP11B1* primer pairs were aligned to the hg38 reference genome using *Minimap2* (12), and the resulting alignment file (BAM format) was sorted and indexed using *SAMtools* (13). SNVs/Indels were subsequently identified using *Clair3* (version 1.0.10) (16).

For the analysis of *CYP21A2*, sequencing reads derived from the A2F/TNXB-R and A2F/TNXA-R primer pairs were first aligned to the reference *CYP21A2-TNXB* sequence using *Minimap2* (12), and then sorted and indexed using *SAMtools* (13). For reads specifically derived from the A2F/TNXA-R amplicon, a target region (positions 4785–12795 in the *CYP21A2-TNXB* reference) was extracted from the BAM file using a custom-developed software tool. Three synthetic marker sequences—C4BF, C4AF and TNXAR, representing gene-specific characteristic regions—were applied to further classify the extracted reads and generated three reads subsets (C4AF/TNXAR, C4BF/TNXAR, and A2F/TNXB-R). The reads were then individually aligned, followed by variant calling and haplotype phasing.

SNVs and indels were identified using *Clair3* (16) and normalized with *Bcftools* (13). Haplotype phasing was performed using *Whatshap* (version 2.2) (17). Identified variants were then annotated based on a pseudogene differential site database, and the number of haplotypes for functional and pseudogene sequences at each site was calculated. Finally, variants were classified as duplication (dup), deletion (del), or non-variant (negative) based on differential marker patterns.

## RESULTS

### The workflow and establishment of NanoCAH

The NanoCAH assay was developed using long-range PCR combined with long-read sequencing on the nanopore sequencing platform, CycloneSEQ G100-ER. A multiplex targeted long-range PCR approach was designed to specifically amplify *CYP21A2*, *CYP21A1P*, and *CYP11B1* (Fig. 1A). The amplicon sizes were optimized to approximately 10 kb, maximizing coverage and resolution of the target genomic regions while maintaining technical feasibility and accuracy in downstream sequencing and analysis.

**Figure 1.**
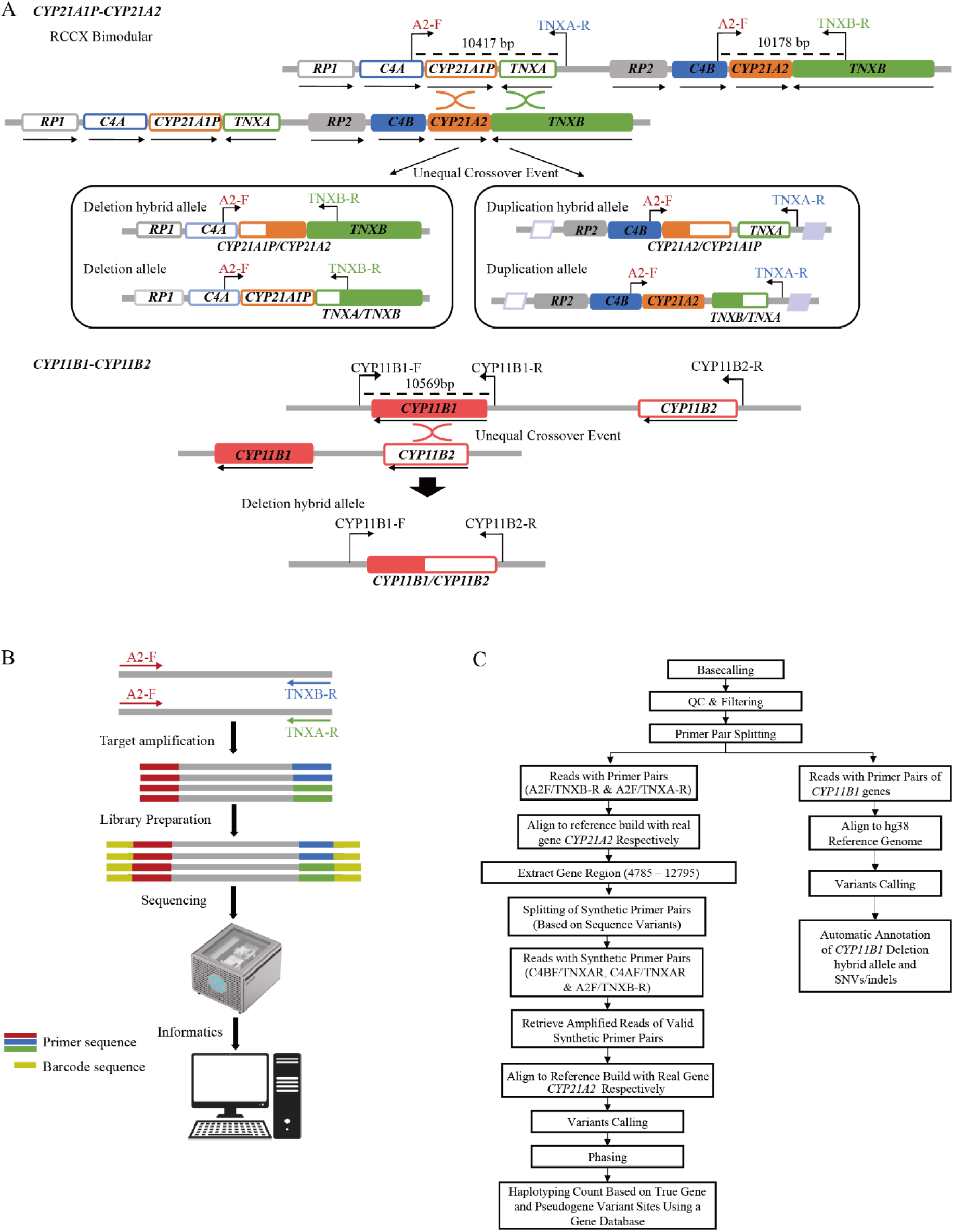
Workflow of NanoCAH assay based on long-read CycloneSEQ sequencing. (A) Design of Long-Range Primers for *CYP21A2* and *CYP11B1*. (B) Workflow for Long-Read Amplification on the CycloneSEQ G100-ER Platform. (C) Customized Bioinformatics Pipeline for CAH Analysis.

For *CYP21A2* and *CYP21A1P*, a forward primer (A2-F) was designed to anneal to the identical sequences upstream of the *C4B* and *C4A* genes, while two specific reverse primers (TNXB-R and TNXA-R) were designed to bind the divergent sequences of the downstream *TNXB* and *TNXA* genes, respectively. The primer pair A2-F and TNXB-R was used to detect SNVs/Indels in *CYP21A2*, as well as minor gene conversions and deletions. In parallel, the A2-F and TNXA-R primer pair was employed to identify duplication alleles. For *CYP11B1*, a primer pair (CYP11B1-F and CYP11B1-R) was designed to flank the *CYP11B1* gene, facilitating the detection of SNVs, INDELs, and minor gene conversions. Additionally, an extra reverse primer (CYP11B2-R) was designed upstream of *CYP11B2* to target potential gene conversion events between *CYP11B2* and *CYP11B1* (Fig. 1A).

The PCR products were subjected to CycloneSEQ library preparation, sequencing and analysis using a customized bioinformatics pipeline designed for LRS CAH (Fig. 1B, C).

### Performance of NanoCAH

A total of 59 samples, comprising 26 singletons and 11 trios, were analyzed using NanoCAH. Conventional methods (MLPA and Sanger sequencing) had previously identified 85 pathogenic variants in these samples, including SNVs/indels, deletions, and duplications. With NanoCAH, we detected 81 variants in *CYP21A2*, consisting of 67 SNVs/indels (83%), 12 deletions (15%), and 2 duplications (2%) (Supplementary Figure 1). These SNVs/indels spanned multiple functional classes, such as nonsense, missense, frameshift, and splice-site variants. The most frequent SNVs/Indels variants of *CYP21A2* were c.518T>A (n = 21), followed by c.293-13C>G (n = 20) and c.1069C>T (n = 7) (Supplementary Figure 2). In addition, NanoCAH detected 14 CNVs, with exon1-7 deletions being the most prevalent (n = 7), followed by exon1-3 deletions (n = 4). Beyond *CYP21A2*, NanoCAH successfully detected 4 pathogenic variants in the two samples carrying *CYP11B1* mutations. Specifically, one carried a heterozygous deletion of Exon 1–6 along with two heterozygous SNVs (c.907delG and c.905A>T), while the other harbored a homozygous c.281C>T variant. Overall, NanoCAH reproduced all 85 variants previously identified by MLPA and Sanger sequencing (100%, 85/85), underscoring its robustness, accuracy, and clinical applicability as a single-platform long-read sequencing approach (Table 1).

**Table 1.**
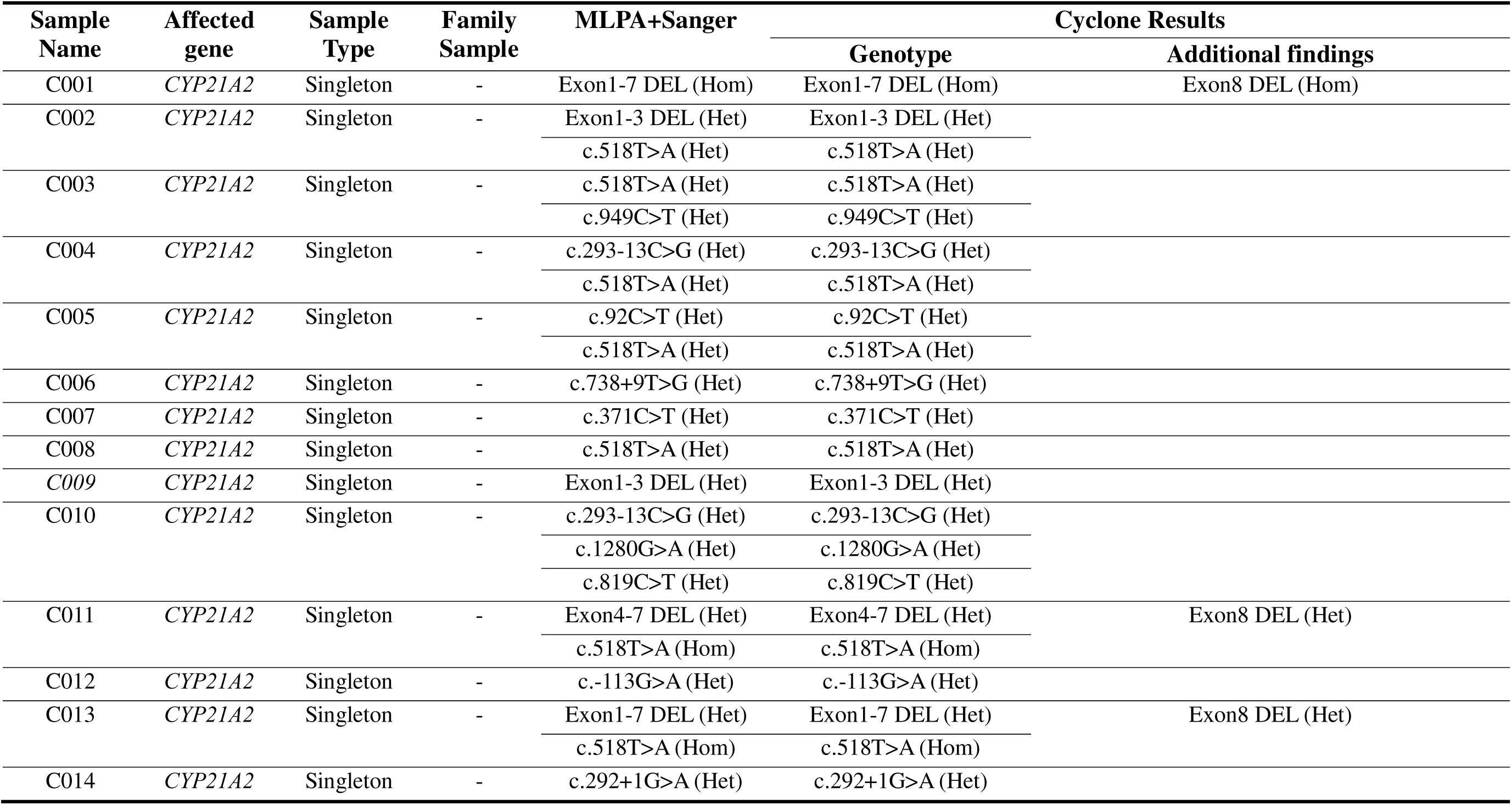
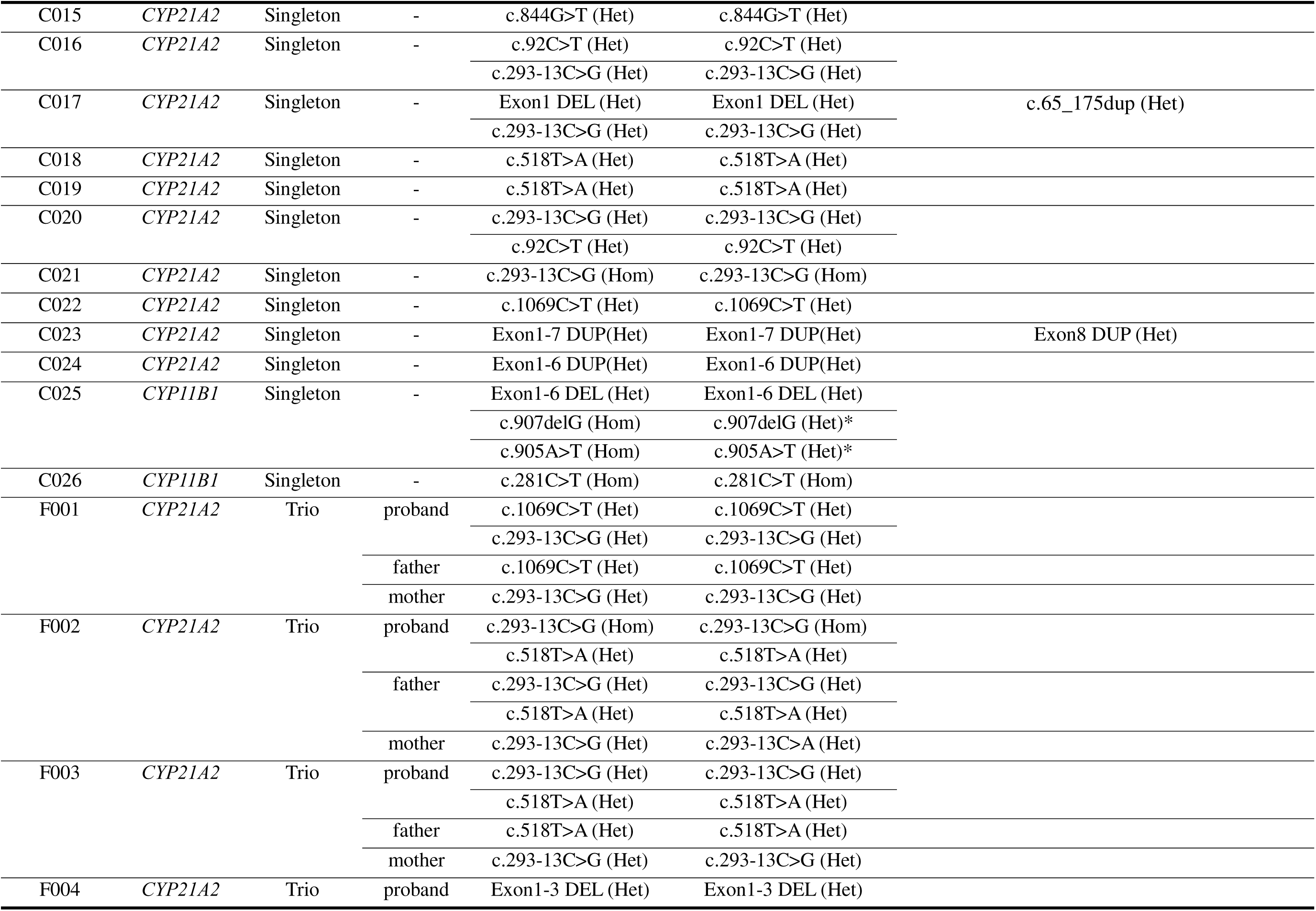
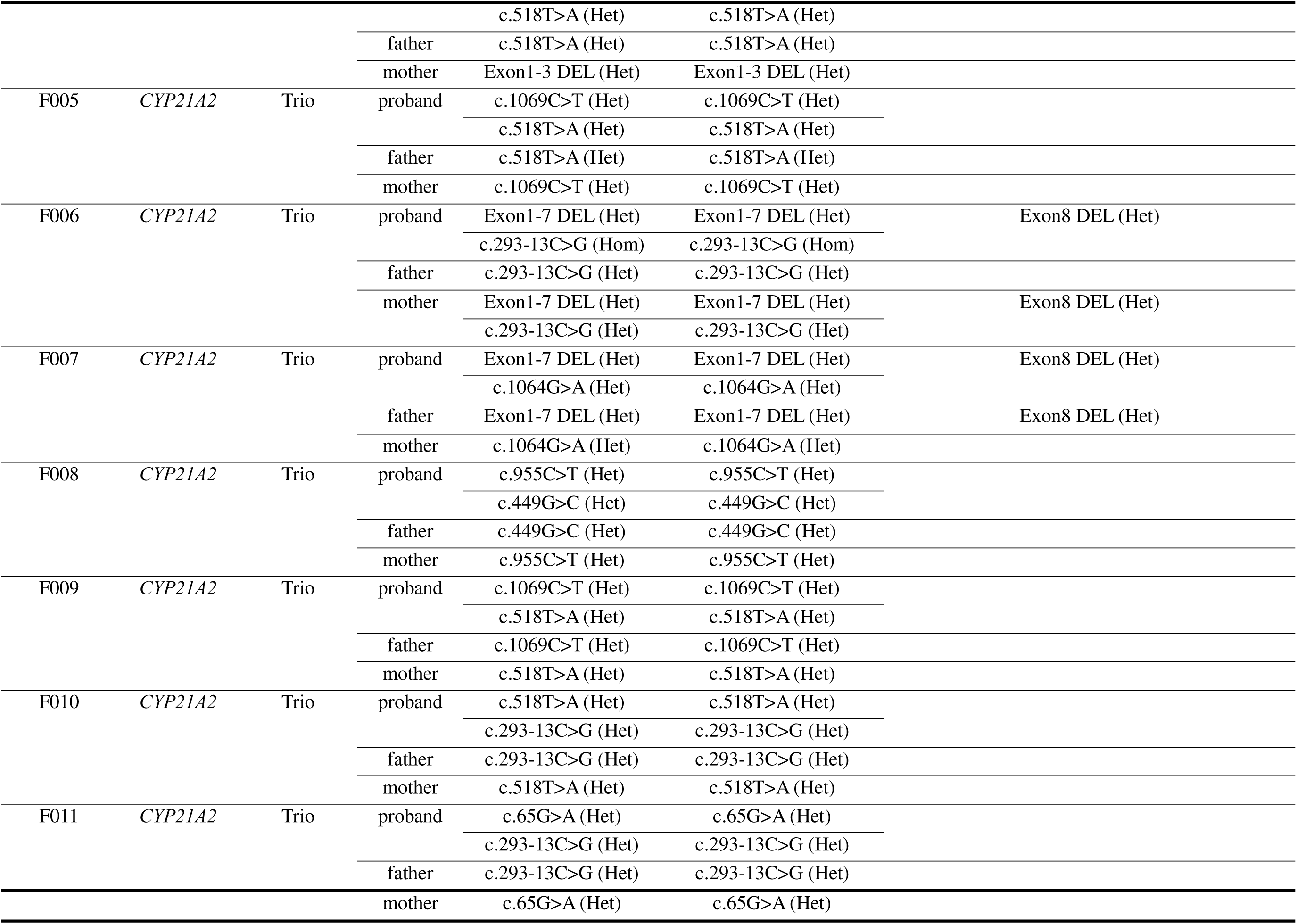
Agreement between Cyclone results and MPLA combined with Sanger sequencing assay for *CYP21A2* and *CYP11B1* genetic testing. DEL, Deletion; Hom, Homozygous; Het, Heterozygous. *For sample C025, the two SNVs from Sanger sequencing appear homozygous. However, due to a structural variant, Sanger only reflects one haplotype. Cyclone results indicate both the structural variant and SNVs, showing heterozygosity when considering both haplotypes. Therefore, the Sanger and Cyclone results are actually consistent.

To further illustrate these capabilities, Figure 2 presents representative variant profiles detected by NanoCAH. These include a heterozygous *CYP21A2* Exon 1–3 deletion (Figure 2A), a compound heterozygote with a c.518T>A SNV and an Exon 1–3 deletion (Figure 2B), and a biallelic SNV configuration involving c.92C>T and c.293-13C>G on separate haplotypes (Figure 2C). Additionally, a *CYP11B1* sample is shown with dual SNVs (c.907delG and c.905A>T) on one haplotype and an Exon 1–6 deletion on the other (Figure 2D). These examples highlight the resolution and phasing power of NanoCAH, particularly in detecting complex variant combinations in regions with high sequence homology.

**Figure 2.**
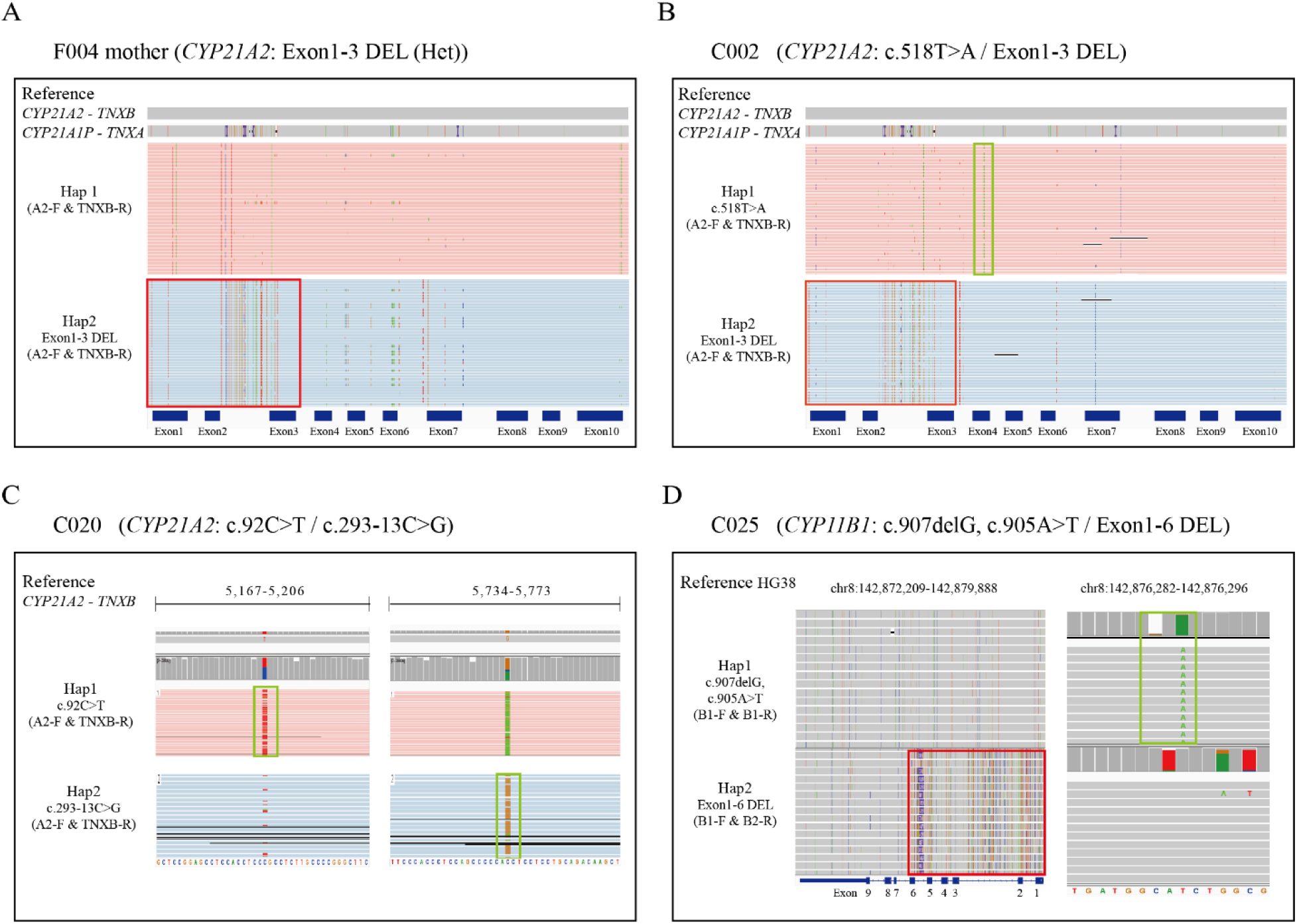
IGV plots show the variations of *CYP21A2* (A-C) and *CYP11B1* (D). In each diagram, the red box indicates the variations caused by pseudogene recombination, specifically the loss of functional gene due to such recombination. For simplicity, this is referred to as a deletion and denoted as DEL. The green box highlights single nucleotide variants (SNVs). Different haplotypes are shown in the upper and lower sections of each plot. (A) Representative sample with heterozygous *CYP21A2* gene deletions; (B) Representative sample with a heterozygous gene deletions and a SNV in *CYP21A2*; (C) Representative sample with pathogenic SNVs in *CYP21A2*; (D) Representative sample with *CYP11B1* gene deletions and pathogenic SNVs/indels.

### Additional Discovery Beyond MLPA and Sanger

A key advantage of NanoCAH is its expanded detection spectrum for deletions and duplications compared to MLPA. One example is the detection of *CYP21A2* Exon 8 deletions/duplications, which are not captured by MLPA probes that span only Exon 1–7. Through full-gene coverage and the detection of two signature SNVs in Exon 8 (c.955C>T and c.1069C>T), as shown in Figure 3, NanoCAH was able to infer deletions/duplications with high confidence. This is exemplified in Figure 3A–D, where MLPA (Figure 3A) detected a heterozygous Exon 1–7 deletion, and Sanger sequencing (Figure 3B) revealed a seemingly homozygous SNV (chr6:32039426 T>A). In contrast, NanoCAH (Figure 3C) identified both the Exon 1–7 deletion and the SNV, and additionally revealed a heterozygous deletion of Exon 8. This finding was independently validated by Sanger sequencing of Exon 8 SNV markers (Figure 3D), verifying the heterozygous state. Across our cohort, NanoCAH identified seven Exon 8 deletions and one Exon 8 duplication, highlighting the expanded diagnostic scope enabled by long-read sequencing (Supplementary Table 1).

**Figure 3.**
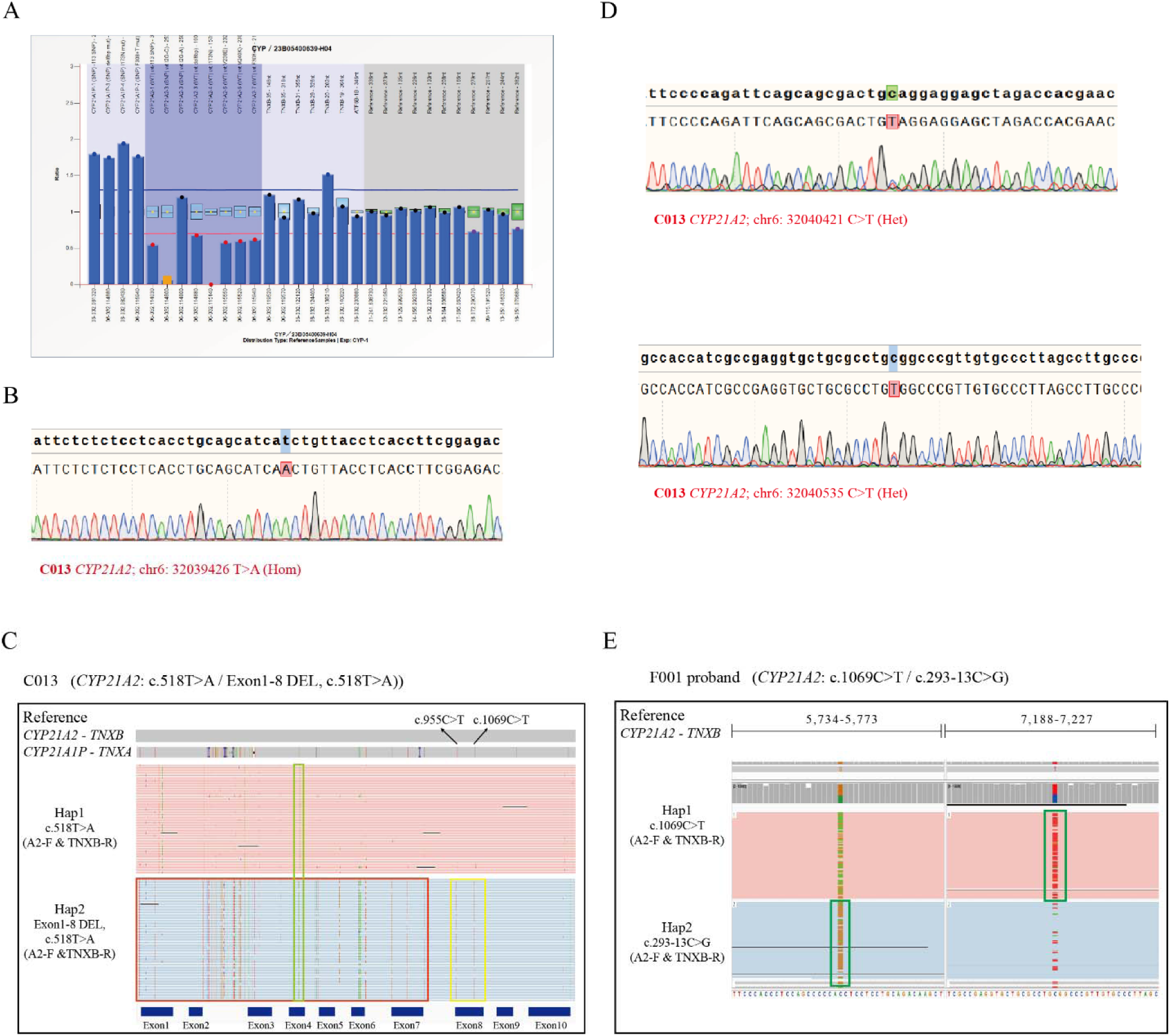
Compared to traditional methods, long-read sequencing enables additional variant detection. (A–C) MLPA, Sanger sequencing, and long-read sequencing results for a sample containing *CYP21A2* gene deletions and a pathogenic SNV. (A) MLPA result shows a heterozygous deletion spanning exon 1–7. (B) Sanger sequencing results reveal a homozygous SNV. (C) Cyclone results not only detect the same deletion (red box) and SNV (green box) but also identify a homozygous deletion of exon 8 (yellow box), which is not detected by MLPA. (D) The additional deletion in exon 8 is validated by Sanger sequencing and confirmed to be present. (E) Long-read sequencing enables the detection of compound heterozygosity (green boxes), which cannot be identified by conventional methods.

In addition, NanoCAH offers precise haplotype-resolved phasing, enabling accurate identification of compound heterozygous configurations. Traditional Sanger-based genotyping relies on trio analysis to resolve *cis*/*trans* configurations. NanoCAH resolves this by directly assigning each variant to its respective haplotype. As shown in Figure 3E, two pathogenic SNVs were mapped to different haplotypes, confirming a compound heterozygous genotype in a single assay. In total, 20 cases were correctly haplotype phased in our study, 18 *in trans* and 2 *in cis* (supplementary Table 2), demonstrating NanoCAH’s effectiveness in resolving compound heterozygous configurations crucial for molecular diagnosis.

Using the NanoCAH long-read workflow, we detected a heterozygous in-frame tandem duplication within *CYP21A2* Exon 1, annotated as NM_000500.9: c.65_175dup (111 bp) (Supplementary Figure 3). Phased long-read alignments showed the head-to-tail junction consistent with a tandem duplication, enabling precise breakpoint localization. Orthogonal confirmation by bidirectional Sanger sequencing across both junctions reproduced the nonreference chimeric sequences (Supplementary Figure 4A): the upstream breakpoint joined the duplicated end to its start (Supplementary Figure 4B), and the downstream breakpoint joined the duplicated start to the downstream reference (Supplementary Figure 4C). Chromatograms at both junctions displayed clean single peaks, concordant with the long-read call. This intragenic 111-bp duplication falls below the effective resolution of routine exon-level CNV assays (e.g. MLPA) and is not reliably captured by standard short-amplicon PCR/Sanger screens. These data illustrate that NanoCAH resolves small in-exon structural variants that conventional methods frequently miss.

## DISCUSSION

Our study demonstrates that LRS enables accurate genotyping of CAH-associated genes with high sequence homology, including *CYP21A2/CYP21A1P* and *CYP11B1/CYP11B2*. In contrast to conventional diagnostic strategies that require MLPA followed by multiple rounds of Sanger sequencing, the CycloneSEQ platform enables simultaneous detection of both SNVs and SVs within a unified workflow, while also providing complete haplotype phasing.

Although CAH is genetically heterogeneous, a majority of clinical cases are caused by mutations in *CYP21A2* and *CYP11B1*, which together account for over 95% of all diagnoses. This study focused on these two genes due to their predominant clinical relevance. Other CAH-related genes are rare and mainly involve SNVs/Indels, which are well-detected by standard short-read sequencing approaches. In contrast, the high sequence similarity between *CYP21A2/CYP21A1P* and *CYP11B1/CYP11B2* complicates accurate analysis with short-read sequencing technologies. Long-read sequencing overcomes this limitation by enabling comprehensive and reliable detection of SNVs/Indels and SVs within these homologous regions.

A key advantage of this approach lies in its ability to identify all the pseudogene-derived clinically relevant SNVs/Indels within regions poorly covered by traditional MLPA assays. For example, the two signature SNVs (c.955C>T and c.1069C>T) in *CYP21A2* Exon 8, undetectable by MLPA due to probe dropout, were reliably captured by NanoCAH assay. Eight samples were further clarified and the gene rearrangement junction sites of the *CYP21A1P*/*CYP21A2* chimera in these samples were defined more precisely. Except for microconversion of SNVs/Indels from *CYP21A1P* to *CYP21A2,* novel rare clinically relevant mutations were also covered by NanoCAH assay. In our cohort, an additional novel 111 bp duplication within Exon 1 of the *CYP21A2* gene, missed by MLPA and Sanger, was identified in sample C017. The duplication was directly visualized in the long-read sequencing data via IGV (Supplementary Figure 3). These findings further illustrate and validate the broader detection spectrum of NanoCAH compared to conventional MLPA and Sanger approaches.

Furthermore, unlike short-read sequencing and Sanger methods which typically require trio-based analysis or statistical inference to resolve haplotype phasing, LRS enables direct haplotyping of compound heterozygous variants within the single proband. This capability is critical for accurate clinical interpretation and genetic counseling, particularly when parental samples are unavailable. In addition, the design of NanoCAH makes it inherently suitable for broader clinical screening purposes. Despite these promising outcomes, one limitation should be acknowledged. This study was involved a limited cohort of 59 positive samples, which, although representative, may not capture the full heterogeneity of CAH-related variants encountered in broader populations. Nonetheless, our findings support the feasibility and clinical potential of the NanoCAH assay for routine genetic testing.

Recent studies using PacBio-based long-read sequencing platforms have demonstrated the high clinical utility of long-read sequencing for CAH. For instance, Yang et al. (2024) analyzed a cohort of 96 CAH patients, showing that targeted LRS improved diagnostic resolution in 14.6% of cases compared to conventional methods, with the ability to uncover novel deletions, duplications, and refine misdiagnosed MLPA results (18). Another newborn screening study in Fujian, China, applied LRS to over 1.7 million newborns, identifying 109 variant alleles across 57 patients with 21-OHD, and estimated a CAH prevalence of 1:26,883 (19). Notably, LRS on nanopore platform demonstrates advantages in cost and clinical practicality. With index, NanoCAH enables multiplexing of samples at a cost of less than $10 per sample—significantly lower than the PacBio-based assays ($20 per sample) (9) and even below MLPA ($16.69 per sample) (20). Furthermore, to achieve comprehensive variant detection, MLPA often requires additional Sanger sequencing and trio-based analysis, bringing the total cost to approximately $50.07 per case (19). In terms of infrastructure, NanoCAH utilizes the compact Cyclone nanopore sequencer, which is more suitable for clinical laboratories than the bulky and costly PacBio Sequel II system. These features make NanoCAH a more practical and scalable solution for routine CAH genotyping in diverse clinical settings.

Looking forward, the streamlined and scalable nature of the NanoCAH workflow makes it well-suited for integration into clinical diagnostic pipelines, particularly in tertiary genetic centers. As sequencing technologies continue to decline in cost and gain in throughput, long-read approaches like CycloneSEQ are expected to become increasingly cost-effective, especially when deployed for batch testing or carrier screening. Moreover, given the recurring challenge of pseudogene interference in many genetic disorders, this workflow can be readily adapted to other complex loci—such as *SMN1*/*SMN2*, *GBA*/*GBAP1*, or *PMS2*/*PMS2CL*—where conventional methods struggle to achieve accurate resolution. Future prospective, multi-center studies with larger and more diverse cohorts will be essential to validate the clinical utility, cost-effectiveness, and scalability of this approach across broader healthcare settings.

## CONCLUSION

In summary, our study demonstrates that long-read sequencing offers a powerful, accurate, and scalable solution for comprehensive genotyping of CAH-associated genes in the presence of complex pseudogene backgrounds. The CycloneSEQ-based NanoCAH workflow not only enables simultaneous detection of SNVs/Indels and SVs in a single assay but also resolves phasing and compound heterozygosity with high confidence. Compared to conventional methods, it improves diagnostic yield, reduces workflow complexity, and captures variants missed by MLPA and Sanger sequencing. These findings support the integration of long-read sequencing into routine molecular diagnostics for CAH and potentially other pseudogene-related disorders. Future large-scale, prospective studies are warranted to validate its clinical utility and cost-effectiveness in broader populations.

## Data Availability

All data produced in the present study are available upon reasonable request to the authors

## Research Funding

This study is supported by the National Key Research and Development Program of China (2023YFC3402504)

## Author Contributions

Shanshan Gu, Guoming Chu, Ruining Cai, and Xiangzhong Sun contributed primarily to manuscript drafting.

Shanshan Gu established the bioinformatics pipeline and performed data analysis.

Guoming Chu was responsible for sample collection.

Ruining Cai and Xiangzhong Sun designed and conducted the experimental work.

Qinglong Lu and Senwen Deng assisted with experimental procedures.

Wanqing Han and Xiaohua Wang contributed to experimental data analysis.

The study was supervised by Jiale Xiang and Rong He, who also contributed to study conception and critical manuscript revision.

All authors reviewed and approved the final version of the manuscript.

