## Supplemental data for "High-Resolution Variant Profiling of CAH-Associated Genes Using a Long-Read Sequencing Assay"

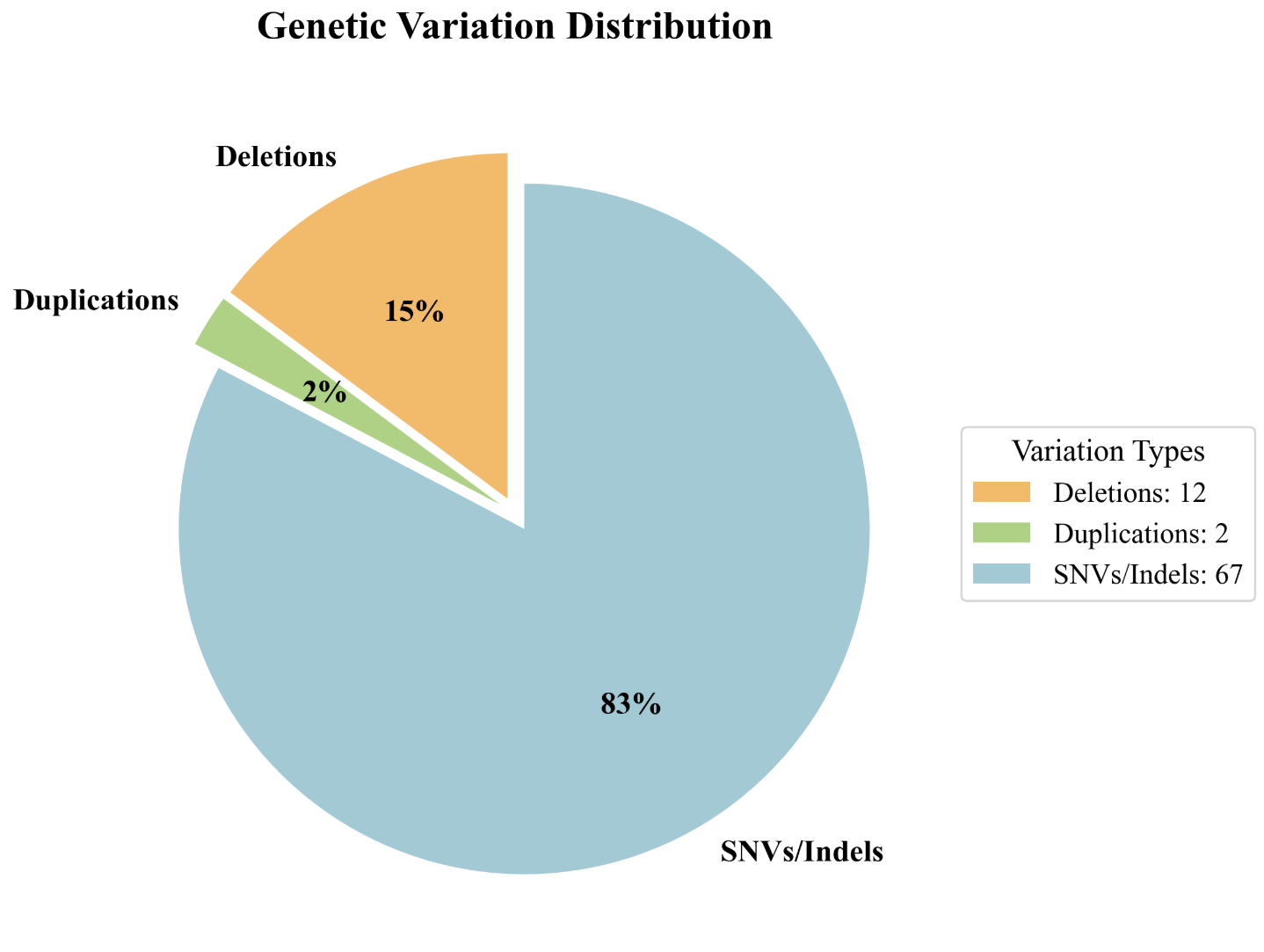

**Supplementary Figure 1. Distribution of Variant Types of *CYP21A2* Identified by NanoCAH.**

A pie chart illustrates the proportion of variant types detected across 57 samples using the NanoCAH workflow. A total of 81 variants were identified: SNVs/indels (83%), deletions (15%) and duplications (2%).

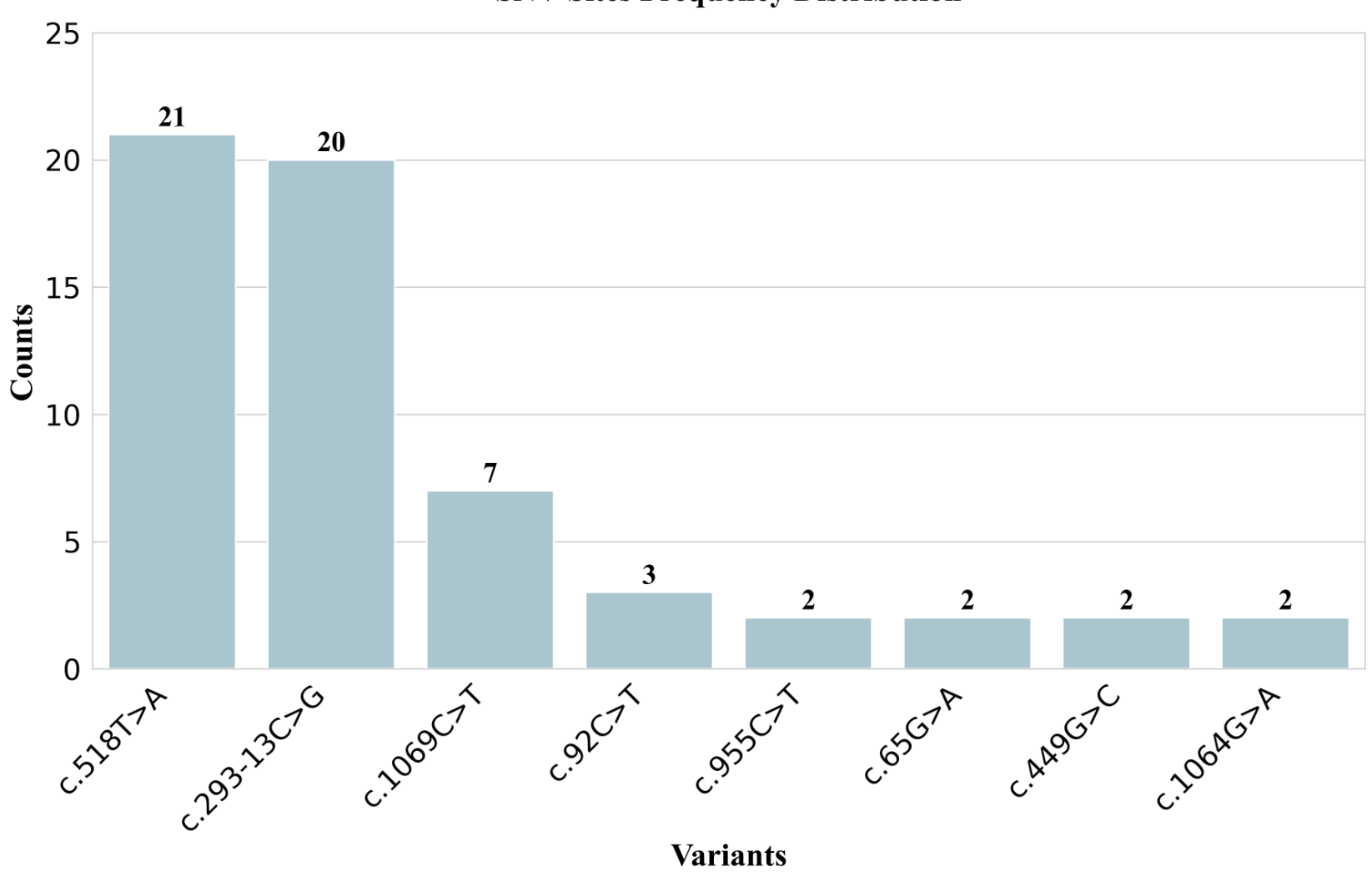

**Supplementary Figure 2. Top 8 Most Frequent SNVs/Indels Identified in *CYP21A2* by NanoCAH.**

The Bar chart shows the top 8 most common SNVs/Indels detected in the *CYP21A2* gene using the NanoCAH workflow. The most common variant is c.518T>A, followed by c.293-13C>G and c.1069C>T. These frequent pathogenic variants highlight known mutational hotspots in *CYP21A2* relevant to CAH diagnosis.

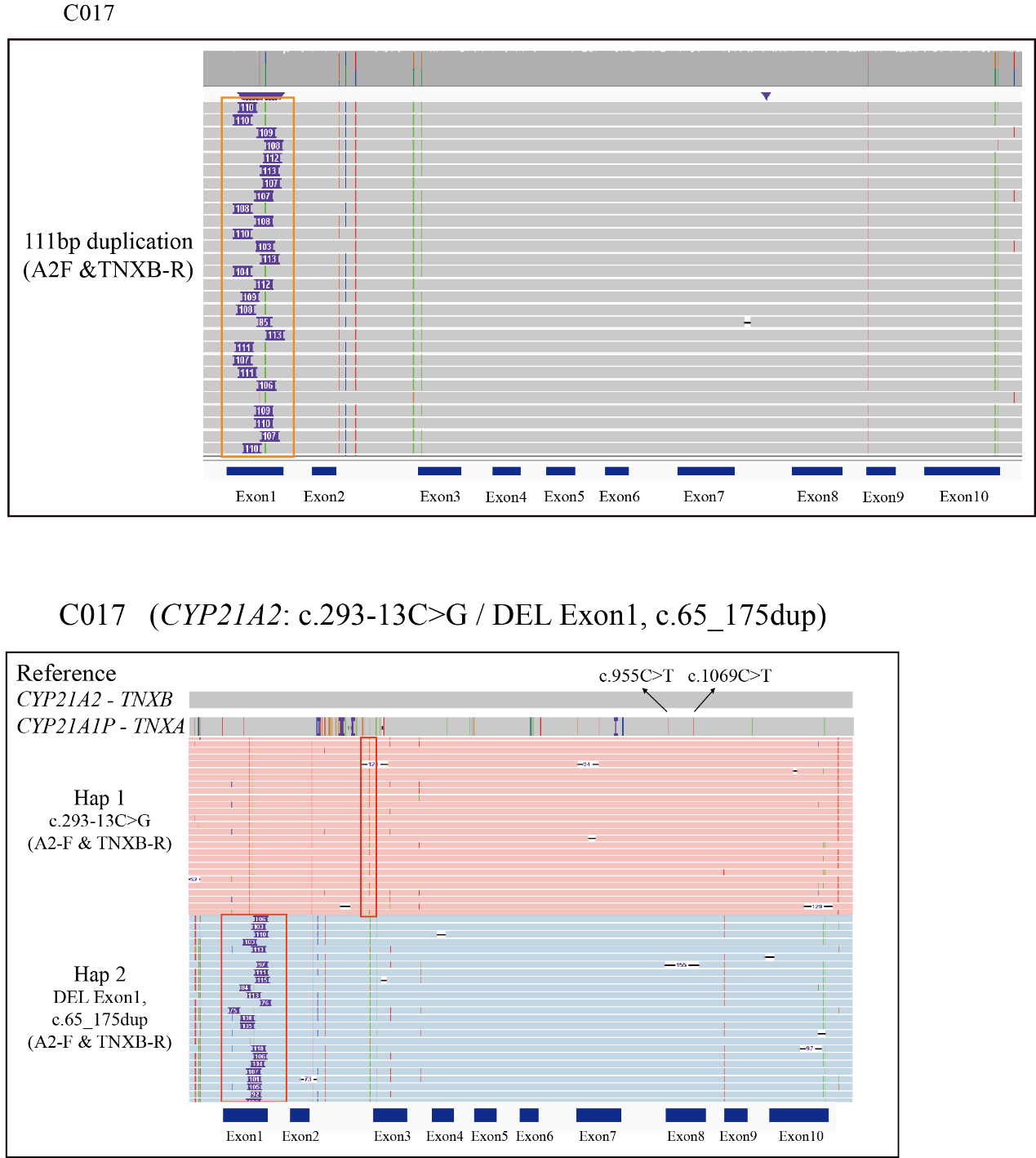

**Supplementary Figure 3. A 111bp Duplication of Exon 1 Detected by NanoCAH.**

Long-read phasing with the NanoCAH workflow reveals two pathogenic alleles *in trans.* Haplotype 1 carries the intronic splice-site variant c.293-13C>G, and haplotype 2 harbours a 111-bp tandem duplication within exon 1 (c.65_175dup).

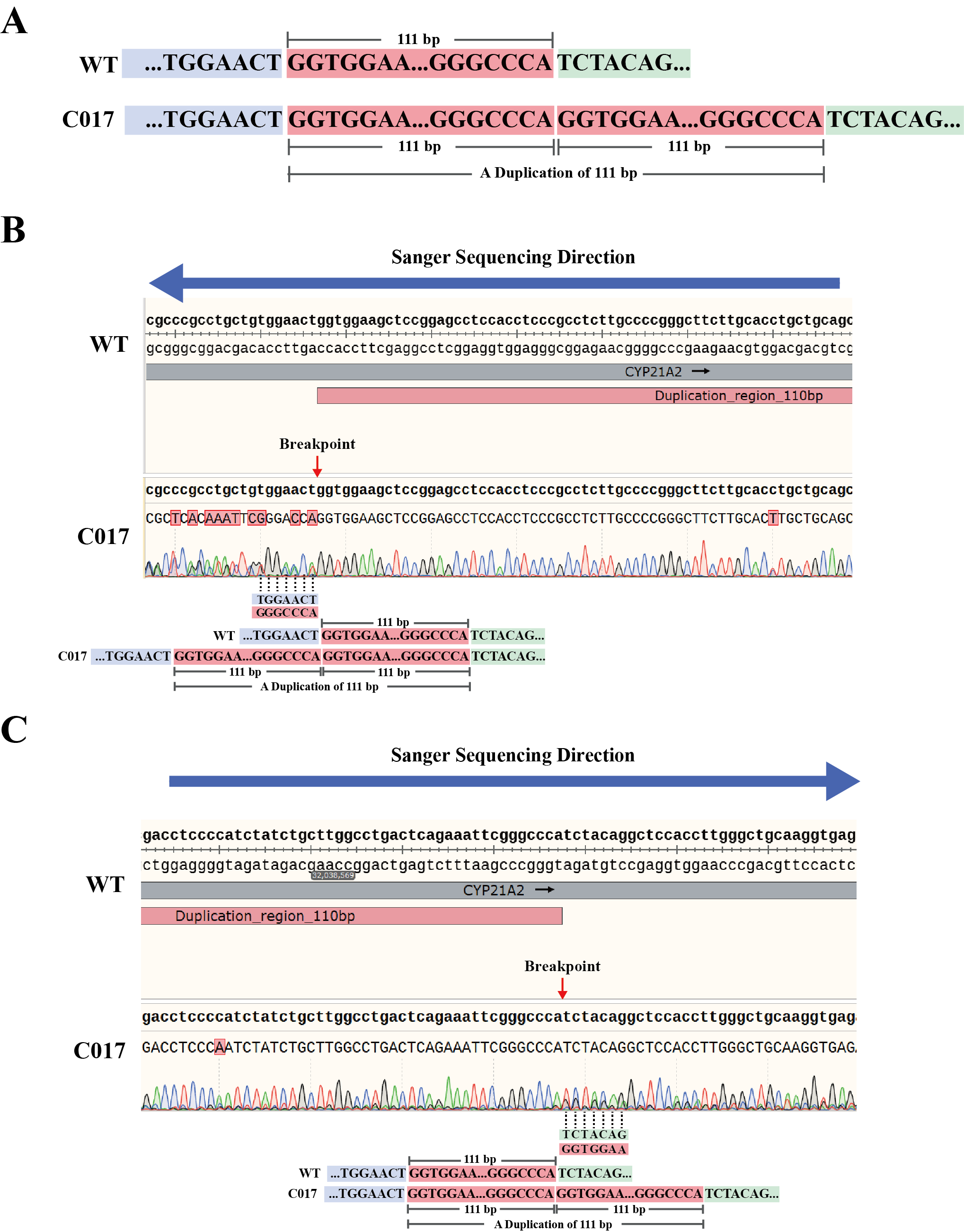

**Supplementary Figure 4. Bidirectional Sanger confirmation of the 111-bp tandem duplication in *CYP21A2* exon 1 (c.65_175dup).**

The red arrow points to the location of the breakpoint. (A) Schematic representation of the 111-bp tandem duplication in *CYP21A2* exon 1 detected in sample C017. (B) Reverse-strand Sanger chromatogram spanning the downstream breakpoint. (C) Forward-strand Sanger chromatogram spanning the upstream breakpoint.

**Supplementary Table 1. Summary of Additional Deletions/Duplications Identified by NanoCAH but Missed by Conventional Methods.**

| **Sample Name** | **Sample Type** | **Family Sample** | **MLPA+Sanger** | **Cyclone Results** | |
| --- | --- | --- | --- | --- | --- |
|  |  |  |  | **Genotype** | **Additional findings** |
| C001 | Singleton | - | Exon1-7 DEL (Hom) | Exon1-7 DEL (Hom) | Exon8 DEL (Hom) |
| C011 | Singleton | - | c.518T>A (Hom) | c.518T>A (Hom) |  |
|  |  |  | Exon4-7 DEL (Het) | Exon4-7 DEL (Het) | Exon8 DEL (Het) |
| C013 | Singleton | - | c.518T>A (Hom) | c.518T>A (Hom) |  |
|  |  |  | Exon1-7 DEL (Het) | Exon1-7 DEL (Het) | Exon8 DEL (Het) |
| C023 | Singleton | - | Exon1-7 DUP(Het) | Exon1-7 DUP(Het) | Exon8 DUP(Het) |
| F006 | Trio | proband | Exon1-7 DEL (Het) | Exon1-7 DEL (Het) | Exon8 DEL (Het) |
|  |  |  | c.293-13C>G (Hom) | c.293-13C>G (Hom) |  |
| F006 | Trio | mother | Exon1-7 DEL (Het) | Exon1-7 DEL (Het) | Exon8 DEL (Het) |
|  |  |  | c.293-13C>G (Het) | c.293-13C>G (Het) |  |
| F007 | Trio | proband | Exon1-7 DEL (Het) | Exon1-7 DEL (Het) | Exon8 DEL (Het) |
|  |  |  | c.1064G>A (Het) | c.1064G>A (Het) |  |
| F007 | Trio | father | Exon1-7 DEL (Het) | Exon1-7 DEL (Het) | Exon8 DEL (Het) |

**Supplementary Table 2.** **Summary of Samples with Multiple Variants and Phasing Configuration.**

| **Sample Name** | **Affected genes** | **Sample Type** | **Family Sample** | **Phasing** | **MLPA+Sanger** | | **Cyclone Results** | |
| --- | --- | --- | --- | --- | --- | --- | --- | --- |
|  |  |  |  |  |  |  | **Genotype** | **Additional detections** |
| C002 | *CYP21A2* | Singleton | - | *in trans* | Exon1-3 DEL (Het) | | Exon1-3 DEL (Het) |  |
|  |  |  |  |  | c.518T>A (Het) | | c.518T>A (Het) |  |
| C003 | *CYP21A2* | Singleton | - | *in trans* | c.518T>A (Het) | | c.518T>A (Het) |  |
|  |  |  |  |  | c.949C>T (Het) | | c.949C>T (Het) |  |
| C004 | *CYP21A2* | Singleton | - | *in trans* | c.293-13C>G (Het) | | c.293-13C>G (Het) |  |
|  |  |  |  |  | c.518T>A (Het) | | c.518T>A (Het) |  |
| C005 | *CYP21A2* | Singleton | - | *in trans* | c.92C>T (Het) | | c.92C>T (Het) |  |
|  |  |  |  |  | c.518T>A (Het) | | c.518T>A (Het) |  |
| C010 | *CYP21A2* | Singleton | - | *in trans* | c.293-13C>G (Het) | | c.293-13C>G (Het) |  |
|  |  |  |  |  | c.1280G>A (Het) | | c.1280G>A (Het) |  |
|  |  |  |  |  | c.819C>T (Het) | | c.819C>T (Het) |  |
| C016 | *CYP21A2* | Singleton | - | *in trans* | c.92C>T (Het) | | c.92C>T (Het) |  |
|  |  |  |  |  | c.293-13C>G (Het) | | c.293-13C>G (Het) |  |
| C017 | *CYP21A2* | Singleton | - | *in trans* | Exon1 DEL (Het) | Exon1 DEL (Het) | | c.65_175dup (Het) |
|  |  |  |  |  | c.293-13C>G (Het) | | c.293-13C>G (Het) |  |
| C020 | *CYP21A2* | Singleton | - | *in trans* | c.293-13C>G (Het) | | c.293-13C>G (Het) |  |
|  |  |  |  |  | c.92C>T (Het) | | c.92C>T (Het) |  |
| C025 | *CYP11B1* | Singleton | - | *in trans* | Exon1-6 DEL (Het) | | Exon1-6 DEL (Het) |  |
|  |  |  |  |  | c.907delG (Hom) | | c.907delG (Het)* |  |
|  |  |  |  |  | c.905A>T (Hom) | | c.905A>T (Het)* |  |
| F001 | *CYP21A2* | Trio | proband | *in trans* | c.1069C>T (Het) | | c.1069C>T (Het) |  |
|  |  |  |  |  | c.293-13C>G (Het) | | c.293-13C>G (Het) |  |
| F002 | *CYP21A2* | Trio | father | *in cis* | c.293-13C>G (Het) | | c.293-13C>G (Het) |  |
|  |  |  |  |  | c.518T>A (Het) | | c.518T>A (Het) |  |
| F003 | *CYP21A2* | Trio | proband | *in trans* | c.293-13C>G (Het) | | c.293-13C>G (Het) |  |
|  |  |  |  |  | c.518T>A (Het) | | c.518T>A (Het) |  |
| F004 | *CYP21A2* | Trio | proband | *in trans* | Exon1-3 DEL (Het) | | Exon1-3 DEL (Het) |  |
|  |  |  |  |  | c.518T>A (Het) | | c.518T>A (Het) |  |
| F005 | *CYP21A2* | Trio | proband | *in trans* | c.1069C>T (Het) | | c.1069C>T (Het) |  |
|  |  |  |  |  | c.518T>A (Het) | | c.518T>A (Het) |  |
| F006 | *CYP21A2* | Trio | mother | *in cis* | Exon1-7 DEL (Het) | | Exon1-7 DEL (Het) | Exon8 DEL (Het) |
|  |  |  |  |  | c.293-13C>G (Het) | | c.293-13C>G (Het) |  |
| F007 | *CYP21A2* | Trio | proband | *in trans* | Exon1-7 DEL (Het) | | Exon1-7 DEL (Het) | Exon8 DEL (Het) |
|  |  |  |  |  | c.1064G>A (Het) | | c.1064G>A (Het) |  |
| F008 | *CYP21A2* | Trio | proband | *in trans* | c.955C>T (Het) | | c.955C>T (Het) |  |
|  |  |  |  |  | c.449G>C (Het) | | c.449G>C (Het) |  |
| F009 | *CYP21A2* | Trio | proband | *in trans* | c.1069C>T (Het) | | c.1069C>T (Het) |  |
|  |  |  |  |  | c.518T>A (Het) | | c.518T>A (Het) |  |
| F010 | *CYP21A2* | Trio | proband | *in trans* | c.518T>A (Het) | | c.518T>A (Het) |  |
|  |  |  |  |  | c.293-13C>G (Het) | | c.293-13C>G (Het) |  |
| F011 | *CYP21A2* | Trio | proband | *in trans* | c.65G>A (Het) | | c.65G>A (Het) |  |
|  |  |  |  |  | c.293-13C>G (Het) | | c.293-13C>G (Het) |  |
